# Diabetes as risk factor to severity of dengue in naïve patients

**DOI:** 10.1101/2024.04.27.24306485

**Authors:** Bárbara F. dos Santos, Flora A. Gandolfi, Bruno H. G. A. Milhim, Fernanda S. Dourado, Gislaine C. D. Silva, Nathalia Zini, Victor Hugo Rezende Gratão, Matheus Pascoal Mariani, Tamires Nasie Abbas, Pedro H. C. Garcia, Rodrigo S. Rocha, Nikos Vasilakis, Maurício L. Nogueira, Cássia F. Estofolete

**Affiliations:** Laboratório de Pesquisas em Virologia (LPV), Faculdade de Medicina de São José do Rio Preto (FAMERP); São José do Rio Preto, São Paulo, Brazil; Department of Pathology, University of Texas Medical Branch; Galveston, Texas, USA; Center for Vector-Borne and Zoonotic Diseases, University of Texas Medical Branch; Galveston, Texas, USA; Institute for Human Infection and Immunity, University of Texas Medical Branch; Galveston, Texas, USA; Hospital de Base (HB), São José do Rio Preto, São Paulo, Brazil

**Author notes:** **Corresponding authors:** Cássia F. Estofolete, M.D., Ph.D.; Maurício L. Nogueira, M.D., Ph.D.

## Abstract

**Background:** Dengue cases can progress to severe ant life-threating forms particularly in subsequent heterologous infections. However, recent studies had explored additional risk factors, including underlying health conditions, even in individuals without prior exposure to dengue, notably, in patients with endothelial dysfunction and chronic inflammation. This study examines the link between diabetes and the development of severe dengue disease in dengue-naive patients during the 2019 dengue outbreak in São Jose do Rio Preto, Brazil.

**Methodology:** We enrolled 529 laboratory-confirmed dengue cases, identified through DENV RT-PCR or NS1 antigen assays in a hospital cohort of acute febrile illness. Subsequently, we investigated the presence of anti-dengue and anti-Zika IgG antibodies. Samples testing positive for Zika were excluded from the analyses. Two groups were analyzed: naïve (DV-), and dengue history (DV+).

**Results:** Initially, presence of diabetes and kidney disease, as well as being dengue-naive, were associated with a higher frequency of severe and potentially severe clinical outcomes. Multivariate analysis identified diabetes as a risk factor, while the presence of anti-dengue antibodies was considered protective. Analysis of dengue naïve samples, highlighted diabetes as an independent risk factor to severe forms of dengue disease. In DV+ patients, no condition was highlighted as a risk factor by univariate analysis or multivariate analysis.

**Conclusions:** We investigated and confirmed diabetes as a risk factor for severe dengue disease in individuals without prior dengue or Zika exposure. Our conclusions raise significant concerns given diabetes’ ever increasing global prevalence and its potential impact on patients with or previous dengue exposure.

**Summary:** The simultaneous escalation of diabetes and dengue worldwide is striking. Notably, diabetes presents as a significant risk factor for severe dengue. This accentuates the necessity of diabetes control in dengue prevention, considering its widespread prevalence and influence on disease severity.

## Introduction

Dengue continues to persist as a significant global public health concern. Recent estimates indicate an annual occurrence of 400 million infections, with approximately 100 million symptomatic and up to 3% fatality rates (1). Over the preceding three decades, the global incidence of dengue has surged by 85.5%, accompanied by a mortality rate increase of 28.07% (2). The reported cases have risen from 30,668,000 in 1990 to 56,879,000 in 2019. Furthermore, DENV remains a significant contributor to worldwide mortality, accounting for 36,055 deaths in 2019 alone (2).

Infection by any of the four distinct DENV serotypes, presents itself as an acute self-limiting febrile illness, that may progress to severe and potentially life-threatening disease with 1-3% mortality rate (3, 4). The pathways leading to the development of severe dengue disease (SDD) are complex and multifaceted, and the interplay between the virus and the host orchestrates the development of severe manifestations manifested as plasma leakage and disrupted hemostasis. A multitude of risk factors have been identified as influential factors in the emergence of severe dengue and mortality.

Secondary heterologous infections by DENV have been linked to severe dengue manifestations (5), attributed to an intense inflammatory reaction and heightened immune response, induced by antibody-dependent enhancement (ADE)(6-9) and resulting in higher viral load, damage to endothelial cells, and leakage of plasma (10). Moreover, a recent study by Katzelnick et al (11) in a pediatric cohort in Nicaragua suggested an increase in dengue severity in children previously exposed to only Zika virus (ZIKV) infection. Although this complex interplay of immune responses between dengue and Zika is not fully understood, it was the first study to show that a recent primary Zika infection followed by dengue infection had a higher probability of symptomatic dengue disease (DEN) compared to those with a history of prior dengue infection, or prior exposure to both viruses, and that the observed outcomes were directly correlated to antibody levels. Similar observations were reported by our group in a Brazilian hospital cohort (12), where DENV-2-symptomatic patients with a previous ZIKV infection, but not DENV, presented with higher frequency of dengue with warning signs (DwS) and SDD, compared to immunologically naïve-patients or those with a history of prior exposure to dengue.

Critically, non-immune elements within the host, including age (13), gender (13, 14), nutritional status (15), genetic background (16) and metabolic disorders such as diabetes (17), have been implicated in the development of SDD. Currently, approximately 537 million people across the globe are affected by diabetes, with nearly 80% (ca. 425 million) concentrated in the Americas, the Middle East & North Africa, Southeast Asia, and the Western Pacific (18). Interestingly, these regions also bear a substantial burden of DEN, as reported by the Global Burden of Disease Study in 2017 (19).

Diabetes is a multifaceted clinical disorder, with type 2 diabetes mellitus (T2DM) representing approximately 95% of all global cases (20). Diabetes is intricately viewed as an endothelial disorder caused by hyperglycemia and if left untreated leads to pan-vascular impairment (21), as well as it is linked to a spectrum of complications, such as diabetic ketoacidosis, diabetic coma, and cardiovascular disease (22). Observational studies in several countries have shown a correlation between diabetes with SDD (23-25). Although the pathogenic mechanism remains unclear, Sekaran et al conducted an extensive review of the role of inflammation in diabetic patients, highlighting the role of inflammatory molecules, such as TNF-a, IL-6, IFN-y, IL-2, NO, and T-helper type 1 cells, endothelial or epithelial cells, and cytoskeleton integrity. These molecules and cells are associated with chronic inflammatory status observed in diabetic patients and are involved in contributing to the development of SDD, especially in endothelial disruption and fluid permeability (26). Lastly, diabetes might contribute to endothelial activation and vascular damage (27).

In this study, we analyzed the association between diabetes, as risk factor, and the occurrence of DWS and SDD in patients who were immunologically naïve to dengue and/or Zika infections. This study took place during the 2019 DENV-2 outbreak in São José do Rio Preto, São Paulo, Brazil.

## Material and Methods

### Ethical considerations

This cross-sectional study was based on data obtained from medical records and disease notification forms for dengue-confirmed patients enrolled in hospital cohort of acute febrile illness between December 2018 and November 2019, in São José do Rio Preto, São Paulo, Brazil. The study was conducted according to the guidelines of the Declaration of Helsinki on retrospective samples collected for routine analysis and surveillance with a waiver of consent approved by the Institutional Review Board of the School of Medicine of São José do Rio Preto (FAMERP) (protocol codes 15461513.5.0000.5415, approved on April 07, 2015, and 14262619.0.0000.5415 on August 13, 2019, respectively). Confidentiality was ensured by anonymizing all samples before data entry and analysis.

### Study area

São José do Rio Preto, located in the state of São Paulo, is an area characterized by hyperendemic dengue transmission [as reviewed in (28)]. During the DENV-2 epidemic of 2019, the city witnessed record-breaking numbers of dengue disease (DEN): 33,120 cases, including 528 with DWS, 29 SDD cases, and 19 deaths (29).

### Sample eligibility for inclusion

This study was conducted using samples from patients with laboratory confirmed DEN, aged 15-years-old and older, and classified according to WHO 2009 (4) and the Brazilian Ministry of Health (30) guidelines. Samples from patients younger than 15-years-old were excluded from this study. After initial care was provided, the suspected cases were reported in the national Notifiable Diseases Information System (SINAN). Blood samples were collected for routine diagnosis and stored at -80°C until they were repurposed for testing for this study. Additional testing also included samples from suspected DENV cases up to 7 days following the onset of symptoms and confirmed with One-Step Real time multiplex DENV PCR using serotype-specific primers and fluorogenic probes described by Johnson et al (31)(Promega, Madison, WI, USA). Samples that tested positive remained in the eligible pool, whereas negative samples were included in the sample pool if they had been tested positive by DENV-NS1 ELISA at the time of hospital admission. Samples with negative DENV RT-PCR and DENV-NS1 ELISA were excluded from subsequent analyses. Lastly, samples were tested by Zika IgG ELISA to detect previous Zika infection (12), and all positive samples were also excluded. Detection of anti-dengue IgM antibodies (ELISA or immunochromatographic testing) was not considered diagnostic confirmation, given the existing possibility of cross-reaction with other flaviviruses (32, 33).

### Molecular methods for dengue investigation

To investigate acute dengue infection, viral RNA was extracted from each clinical sample using the QIAamp Viral RNAMini kit (QIAGEN^®^, Germantown, MD, USA), following the manufacturer’s instructions. Viral RNA was analyzed for the presence of DENV according to the protocol described by Johnson et al. (31). One-Step Real time multiplex PCR assays were performed using the GoTaq® Kit (Promega, Madison, WI, USA). In fourplex reaction mixtures, 50 pmol each of DENV-1- and DENV-3-specific primers, 25 pmol each of DENV-2- and DENV-4-specific primers, and 9 pmol of each probe were combined in a 50-µL volume total reaction mixture. Real-time PCR was performed on a 96-well plate and analyzed on the QuantStudio™ Dx instrument (Thermo Fisher Scientific, Waltham, MA, USA) with the following conditions: 50 ◦C for 30 m, followed by 45 cycles of 95 ◦C for 15 s, and 60 ◦C for 1 min. The results were interpreted as positive when Ct values were less than or equal to 37 (31).

### Investigation of previous dengue or Zika infection

Samples from patients with confirmed DENV infection were evaluated for history of DENV and ZIKV infection using an enzyme-linked immunosorbent assay (ELISA). Prior infection with Zika infection was evaluated as described previously (12). Briefly, samples were screened using the Panbio® dengue IgG Capture ELISA (Abbot^TM^, Illinois, USA), EUROIMMUN human anti-ZIKV IgG ELISA kit (EUROIMMUN, EURO-AG, Luebeck, Germany). All assays were performed according to the manufacturer’s recommendations using the corresponding positive and negative controls. ELISA plates were read using a Spectramax Plus ELISA reader at 450 nm (Molecular Devices, LLC, San Jose, CA, USA). Results were expressed in Standard Units (PU) and interpreted as <0.8 negative, >0.8 and <1.1 equivocal and >1.1 positive. For statistical analyses two groups of samples were defined: (i) dengue negative (DV-), and (ii) dengue positive (DV+).

### Demographic and clinical characteristics

Demographic and clinical data such as sex, age, comorbidities, and clinical observations to classify the severity of dengue were obtained from the participants’ electronic medical records or SINAN reporting forms. According to 2009 WHO (4) and the Brazilian Ministry of Health (30) guidelines, DEN was classified as: (i) dengue without warning signs (DwWS), (ii) dengue with warning signs (DWS), or (iii) severe dengue disease (SDD).

### Statistical analysis

Firstly, the descriptive statistics were calculated to summarize the demographic characteristics of the samples. Next, adherence chi-square test was conducted to assess the difference among variable frequencies into the groups. Additionally, the data were analyzed for normality distribution using the Shapiro-Wilk test. ANOVA with the Bonferroni post-hoc test was used to evaluate the significance of differences between each group in normally distributed data. When normality was not met, the Kruskal-Wallis test was used to determine whether there was a difference between the means of the groups. Pearson’s chi-square test was used to determine whether the expected frequency in the groups was met. Finally, binary logistic regression was performed to verify the predictors of severe forms (dengue with warning signal or severe dengue) after meeting the prerequisite of not exhibiting multicollinearity, with tolerance value > 0,1 and variance inflation factor (VIF) < 10. The variables that were significant in univariate analysis were subjected to multivariate analysis, with significance defined as p<0.05. A 0.05 (5%) significance level was adopted for all tests, and all data were tabulated and analyzed with SPSS software for IOS (version 28, SPSS, Inc; Chicago, Il, USA).

## Results

### General characteristics of the study population

Between December 2018 and November 2019, 2,990 serum samples from patients with suspected dengue were obtained, of which 1,490 were laboratory confirmed using DENV-specific RT-PCR or dengue NS1 ELISA assays. Of these, 529 were eligible based on the availability of medical records to classify the severity of the clinical presentation. The majority of the samples were collected from females (52.36%; 277/529; p < 0.27), adults (15-59 years-old) (86.61%; 453/523; p < 0.001) and classified as dengue without warning signs (86.21%; 504/529; p < 0.001). Samples were classified into two groups, based on their serological status: (1) naïve to DENV (DV-), n=96 (18.15%); and (2) DV+, n=433 (81.85%) (p < 0.001) (**Fig. 1**).

**Figure 1.**
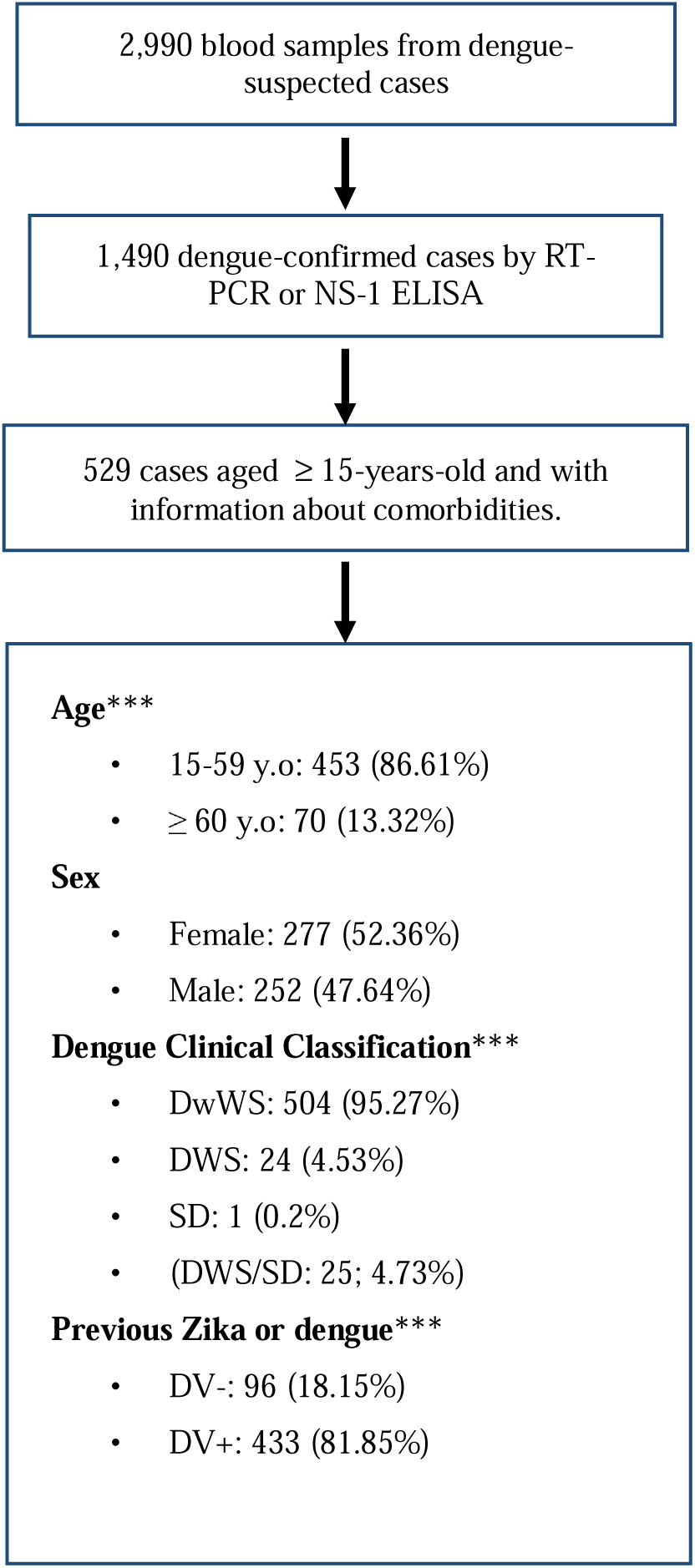
Flowchart of eligibility criteria and general characteristics of enrolled patients’ samples.

### Association of demographic characteristics, comorbidities, and severity of dengue

Given the low number of SDD, severe cases (DwS and SDD) were combined as a single group: DWS/SD. Comparison of DwWS and DWS/SDD showed no statistical differences in regard to age (p = 0.062), or gender (p = 0.109). A greater frequency of severe cases was observed among DV-individuals (p < 0.001). Critically, a higher frequency of individuals with DWS/SD had diabetes (p < 0.011), and kidney disease (p = 0.047) **(Table 1).**

**Table 1.** Demographic and serological characteristics and comorbidities according with severity of dengue.

|  | Severity (DWS or SD) |  |  |  |  |  | p value<sup>1</sup> |
| --- | --- | --- | --- | --- | --- | --- | --- |
|  | DwWS |  |  | DWS/SD |  |  |  |
|  | N response | N positive | % | N response | N positive | % |  |
| **Age group** |  |  |  |  |  |  |  |
| Adult | 498 | 435 | 87,35% | 25 | 18 | 72,00% | 0.062 |
| > 59 yo | 498 | 63 | 12,65% | 25 | 7 | 28,00% |  |
| **Gender** |  |  |  |  |  |  |  |
| Female | 504 | 260 | 51,59% | 25 | 17 | 68,00% | 0.109 |
| Male | 504 | 244 | 48,41% | 25 | 8 | 32,00% |  |
| **Comorbidities** |  |  |  |  |  |  |  |
| Diabetes | 504 | 33 | 6,55% | 25 | 5 | 20,00% | **0.027** |
| Liver disease | 504 | 2 | 0,40% | 25 | 0 | 0,00% | 1 |
| high blood pressure | 504 | 64 | 12,70% | 25 | 5 | 20,00% | 0.354 |
| Peptic disease | 496 | 2 | 0,40% | 25 | 0 | 0,00% | 1 |
| Auto immune disease | 504 | 3 | 0,60% | 25 | 0 | 0,00% | 1 |
| Kidney disease | 504 | 0 | 0,00% | 25 | 1 | 4,00% | **0.047** |
| **Previous infection** |  |  |  |  |  |  |  |
| anti-DV IgG negative | 504 | 83 | 16,47% | 25 | 13 | 52,00% | **<0.001** |
| anti-DV IgG positive | 504 | 421 | 83,53% | 25 | 12 | 48,00% |  |

<sup>1</sup>Chi-square test or Exact Fisher's Test, significant value: $< 0.05$ , confidence interval 95%.
y.o.: years old; DwWS: dengue without warning signs; DWS: dengue with warning signs; SD: severe dengue; DV: dengue

Univariate analysis showed that individuals older than 59-years-old presented a higher risk (OR 2.685; 95% CI 1.079-6.685; p = 0.034) to progress to severe DEN (DWS or SDD) when compared to control (individuals aged 59 or younger). Similarly, diabetes was also linked to a higher risk (95% CI 1.259-10.111; p = 0.017) for progression to severe forms of DEN. Previous exposure to dengue was also not related to severe forms of DEN in univariate analysis. In fact, the occurrence of previous dengue infection was associated with a protective effect (OR 0.182; 95% CI 0.080-0.413; p < 0.001) similar to what was described before (12) **(Table 2).** The multivariate analysis showed that diabetes was a contributing risk factor for progression to SDD (OR 4.805; 95% CI 1.543-14.961; p = 0.007), while again prior exposure to dengue (OR 0.138; 95% CI 0.058-0.329; p < 0.001) and, now, gender were protective (OR 0.386; 95% CI 0.158-0.946; p = 0.037).

**Table 2.** Risk of severe forms of dengue according with characteristics of enrolled individuals.

|  | Univariate analysis<sup>1</sup> |  |  |  |
| --- | --- | --- | --- | --- |
|  | p value | OR | LL | UP |
| **Age group** |  |  |  |  |
| Adult | - | 1 | - | - |
| > 59 y.o. | **0.034** | 2.685 | 1.079 | 6.685 |
| **Gender** |  |  |  |  |
| Female | - | 1 | - | - |
| Male | 0.115 | 0.501 | 0.213 | 1.183 |
| **Comorbidities** |  |  |  |  |
| Diabetes | **0.017** | 3.568 | 1.259 | 10.111 |
| Liver disease | NA | - | - | - |
| High blood pressure | 0.295 | 1.719 | 0.623 | 4.740 |
| Peptic disease | NA | - | - | - |
| Autoimmune disease | NA | - | - | - |
| Kidney disease | NA | - | - | - |
| **Previous infection** |  |  |  |  |
| anti-DV IgG negative | - | 1 | - | - |
| anti-DV IgG positive | **< 0.001** | 0.182 | 0.080 | 0.413 |
| **Multivariate analysis<sup>1</sup>** |  |  |  |  |
| Diabetes | **0.007** | 4.805 | 1.543 | 14.961 |
| anti-DV IgG positive | **< 0.001** | 0.138 | 0.058 | 0.329 |
| Gender | **0.037** | 0.386 | 0.158 | 0.946 |

<sup>1</sup> Binary logistic regression; OR: odds ratio; LL: low limit; UP: upper limit
y.o.: years old; DwWS: dengue without warning signs; DWS: dengue with warning signs; SDD: severe dengue disease; DENV: dengue virus

Univariate analysis of samples from naïve-patients (DV-), revealed a higher risk (6.844; 95% CI 1.550-30.220; p = 0.011) for progression to severe forms of DEN (DWS or SDD) in individuals older than 59-years old, and a risk of 24.60 (95% CI 2.331-259.619; p = 0.008) in diabetics. Being male showed a protective effect (OR 0.252; 95% CI 0.071-0.887; p = 0.032). When all variables were analyzed concurrently through multivariate analysis, only diabetes showed to be a risk factor, with an OR of 24.300 (95% CI 2.302-256.48; p = 0.008). **(Table 3).**

**Table 3.**
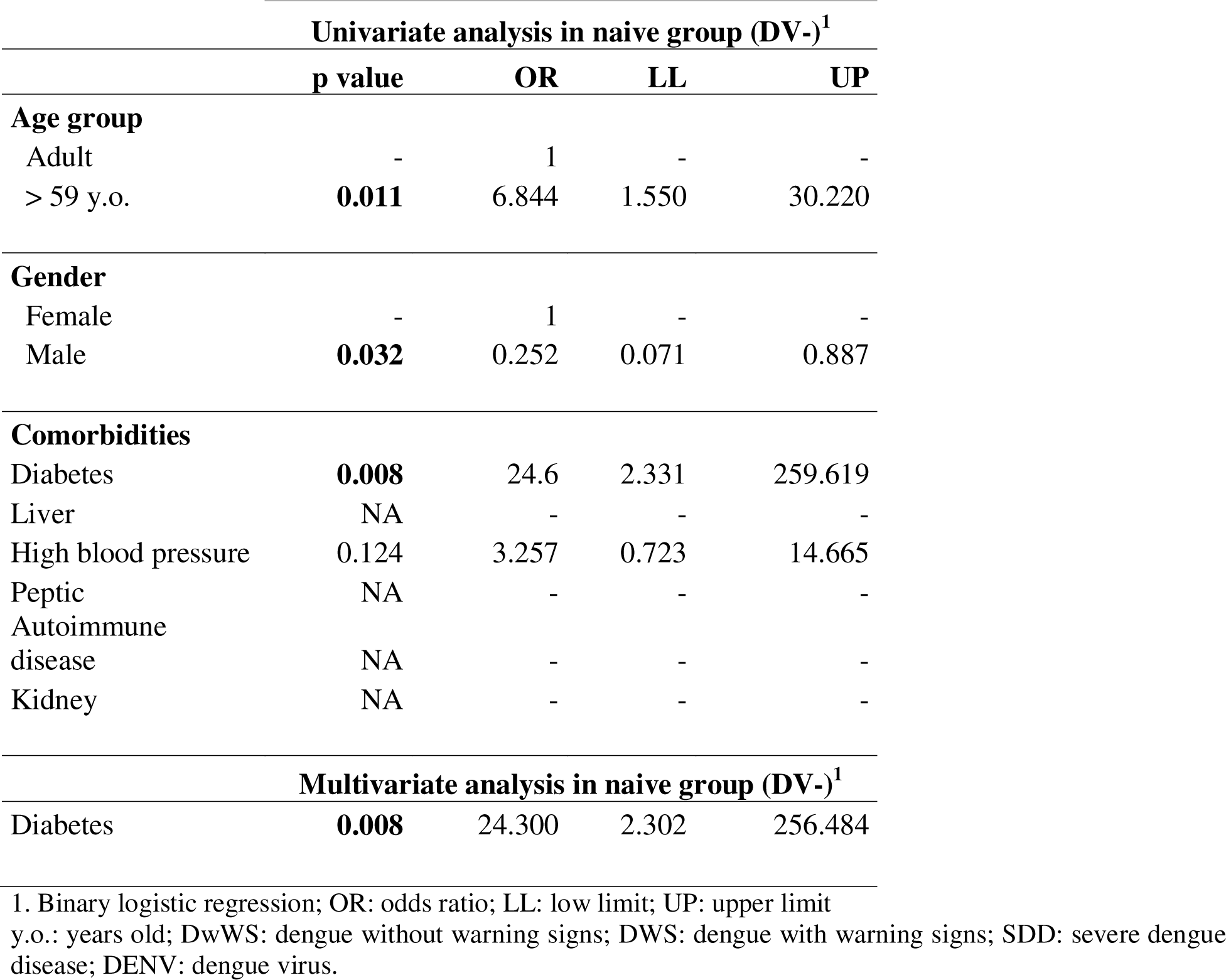
Univariate and multivariate analysis evaluating risk of severe forms of dengue according with characteristics of individuals.

On the other hand, in samples from patients with a history of previous dengue infection (DV+), no conditions were related to risk to severe forms in uni or multivariate analysis.

**Univariate analysis in positive group (DV+)<sup>3</sup>**
|  | p value | OR | LL | UP |
| --- | --- | --- | --- | --- |
| **Age group** |  |  |  |  |
| Adult | - | 1 | - | - |
| > 59 y.o. | 0.290 | 2.057 | 0.541 | 7.824 |
| **Gender** |  |  |  |  |
| Female | - | 1 | - | - |
| Male | 0.413 | 0.602 | 0.179 | 2.030 |
| **Comorbidities** |  |  |  |  |
| Diabetes | 0.264 | 2.431 | 0.511 | 11.574 |
| Liver | NA | - | - | - |
| High blood pressure | 0.756 | 1.277 | 0.273 | 5.979 |
| Peptic | NA | - | - | - |
| Autoimmune disease | NA | - | - | - |
| Kidney | NA | - | - | - |
| **Multivariate analysis in positive group (DV+)<sup>3</sup>** |  |  |  |  |
|  | NA | - | - | - |

1. Binary logistic regression; OR: odds ratio; LL: low limit; UP: upper limit y.o.: years old; DwWS: dengue without warning signs; DWS: dengue with warning signs; SDD: severe dengue disease; DENV: dengue virus

## Discussion

In this study, we observed the influence of diabetes as an independent risk factor for progression to severe forms of DEN (DWS or SDD) in DV-individuals. This is an alarming observation as the global prevalence of diabetes is dramatically increasing (18). The World Health Organization (WHO) in its 2023 World Health Statistics annual report, highlighted the alarming impact of noncommunicable diseases (NCDs) (among them diabetes), which contributed up to 74% of all global deaths and 63% of global disability-adjusted life-years (DALY’s) in 2019 alone. Among the leading causes of death, four stand out: cardiovascular disease, cancer, chronic respiratory disease, and diabetes, responsible for 33.3 million deaths in 2019 (34). Diabetes, which was directly responsible for 2 million [1.4-2.7 million] deaths, may require further attention as it elevated association with cardiovascular (35), including heart (coronary artery disease, angina pectoris, heart failure), brain (stroke, encephalopathy), kidney (diabetic kidney disease) and ocular (retinopathy) disorders (21, 36).

Recently available data from the International Diabetes Federation (IDF) also underscore the staggering impact of diabetes, revealing that 537 million people were afflicted in 2021. Projections anticipate a 12.2% surge, with the global diabetic populace escalating to 783 million by 2045 (18). Among our collected samples, diabetes was the predominant comorbidity documented, closely followed by hypertension. These two conditions are components of a significant syndrome known as metabolic syndrome, the ultimate consequences of which can encompass cardiovascular diseases and potential fatality if not managed effectively. Brazil is rank as the country in South America with the highest number of people with diabetes among adults (20 – 79-year-old age group, with a total of 15.7 millions of cases as of 2021 (18).

From a pathophysiological standpoint, both severe forms of DEN (DWS or SDD), can be seen as an heightened immune response affecting the hemodynamic stability of the body, resulting in intravascular fluid extravasation, hemoconcentration, accumulation of fluids in the extravascular space, as well as potential bleeding and involvement of vital organs, including liver, respiratory system, and central nervous system (4). Furthermore, the presence of comorbidities in DENV-infected patients should be considered a warning sign for increased attention to the patient by the health authorities, including conducting a complete blood count, and a more rigorous monitoring of symptoms (4, 30).

Age is already considered a risk factor for the development of severe forms of dengue disease (20). In a comprehensive systematic review and meta-analysis conducted between 2008 and 2020, which included over 80% of the studies from Asia and Latin America, Sangkaew et al. (6) highlighted diabetes [OR 4.38 (2.58-7.43)], hypertension [OR 2.19 (1.36-3.53)], renal disease [OR 4.67 (2.21-9.88)], and cardiovascular disease [OR 2.79 (1.04-7.50)] as preexisting conditions associated with SDD. In a similar retrospective case-control study conducted between 2005 and 2008 in Singapore (21), the presence of two or more comorbidities was associated with a higher risk of SDD in patients older than 60 years. When diabetes coexisted with a cardiac disorder the risk was four times higher [8.02; 1.40-45.92] compared to diabetes alone [2.21; 1.10-5.52]. The risk was also increased when diabetes coexisted with hypertension [2.68; 1.07-6.68], hyperlipidemia [4.25; 1.34-13.52], or cardiac disorders with hyperlipidemia [5.79; 1.03-32.64]. Thus, in addition to serving as a significant risk factor for the development of severe forms of dengue on its own, diabetes may also be associated with the other cited risk factor, such as obesity, renal disease, hypertension.

In diabetes, the occurrence of hyperglycemia stems from tissues displaying reduced sensitivity to insulin, which triggers an increased release from pancreatic beta-cells. This continuous secretion eventually culminates in pancreatic fatigue and irreversible impairment (37). The disrupted insulin action and secretion that give rise to increased blood glucose levels can upset the equilibrium between nitric oxide and reactive oxygen species in endothelial cells. Nitric oxide’s crucial role in upholding vascular balance and promoting dilation is significant (38).

Oxidative stress and reactive oxygen species have the potential to render nitric oxide inactive due to the production of endothelin 1, culminating in vasoconstriction and platelet aggregation (39). Moreover, these elements can trigger pathways associated with inflammation, marked by the increased expression of monocyte chemoattractant protein-1 (MCP-1), selectins, vascular cell adhesion molecule-1 (VCAM-1), and intracellular cell adhesion molecule-1 (ICAM-1). This sequence involves monocyte adhesion to the endothelium, activation of macrophages, and heightened secretion of interleukin-1 (IL-1) and TNF-alpha, thus amplifying the inflammatory response (40). Vasoconstrictive agents are additionally produced through an escalated cyclooxygenase-2 (COX-2) pathway, characterized by elevated thromboxane A2 levels and reduced prostacyclin production. This further adds to the disruption of vascular control and impairment of endothelial function (40). This intricate cascade forms the foundation for the prevalent vascular events observed in diabetic patients. As endothelial dysfunction is induced by pro-inflammatory factors that foster vasoconstriction and a prothrombotic state, it becomes plausible to predict the onset of vascular events.

Thus, T2DM can be conceptualized as an endothelial disorder, giving rise to a comprehensive vascular condition that ultimately leads to a wide spectrum of micro- and macrovascular complications affecting vital organs such as the heart, brain, kidneys, and eyes (21). The mechanism by which heart disease leads to organ dysfunction and more severe forms of dengue is not well understood. It is hypothesized that heart disease resulting in reduced cardiac output, in the face of the demand caused by viral infection, may lead to insufficient renal blood flow, which is highly regulated by oxygen levels. The result would be ischemia, toxic injury to the kidneys, and subsequent systemic effects (41, 42). Besides, the reduced clearance of inflammatory cytokines due to chronic or acute kidney damage would be a potential mechanism contributing to the severity of dengue (27). The lack of differentiation between the types of diabetes was a limitation found in our study. However, it is important to consider that endothelial dysfunction is a hallmark of the disease, encompassing its various nuances.

While the presence of diabetes has been recognized as a potential enhancer for severe forms of dengue, an emerging trend in the literature highlights an increasingly reported occurrence: cases of diabetes decompensation, particularly in the form of diabetic ketoacidosis, when coexisting with dengue. Although these reports are predominantly confined to case studies, they underscore substantial concerns pertaining to diabetes management in the context of infection, which is the main precipitating factor for complication such as ketoacidosis (43-45).

Generally, the DV+ group is defined as patients with a prior documented exposure to dengue thus susceptible to a higher risk of progression to SDD due to the influence of antigenic enhancement (ADE). However, in our study, the DV+ group did not exclusively consist of secondary dengue cases commonly associated with ADE but rather encompassed reports of multiple (multitypic) dengue infections. This is in line with the hyperendemic nature of dengue circulation in Sao Jose do Rio Preto, where over 70% of the city’s population has already been infected multiple times with various dengue serotypes (28), confirming the reduced risk of disease seen in multitypic DENV infections observed in other locations (46-48).

The development of SDD is a complex and multifaceted process primarily seen in cases of secondary heterologous DENV infections. Such infections can lead to various outcomes, including a heightened inflammatory response that results in elevated viral levels and intensified immune activation (6-9). However, observations of severe forms occurring in individuals without prior dengue infections highlight an even greater complexity in understanding the pathogenesis of such disease forms. Diabetes, a pan-endothelial, pro-inflammatory disease with a high prevalence in the population, poses a challenge in addressing dengue, especially due to the spatial overlap of the two conditions. Recognizing potential risk factors, as demonstrated in this study, is the first step in shaping therapeutic strategies to reduce the potential morbidity and mortality associated with the coexistence of these diseases.

## Data Availability

The dataset from this study is available on the Mendeley Data platform and may be accessed through the following Doi: 10.17632/f8fhyk9ff6.1

## Acknowledgements

We acknowledge the Laboratório Multiusuário (LMU) at Faculdade de Medicina de São José do Rio Preto (FAMERP) for the availability of the MiSeq System (Illumina). This research was funded by the Centers for Research in Emerging Infectious Diseases “The Coordinating Research on Emerging Arboviral Threats Encompassing the Neotropics (CREATE-NEO)” grant 1U01AI151807 by the National Institutes of Health (NIH/USA) to NV, by INCT Dengue Program grant 465425/2014-3, by INCT Viral Genomic Surveillance and One Health grant 405786/2022-0, and by the São Paulo Research Foundation (FAPESP) via grant 2022/03645-1 awarded to MLN and 2022/09229-0 to CFE. MLN is a CNPq Research Fellow. The author’s contributions: BFS, NV, MLN, CFE conceived and designed the study. BHGAM, FSD, GCDS, NZ performed diagnosis. BFS, FAG, VHRG, MPM, TNA, PHCG, RSS performed the collection of data from medical reports. BFS and BHGAM performed the data analysis, and interpretation. BFS wrote the first draft of the manuscript. BFS, NV, MLN, CFE edited and revised the manuscript. NV, MLN, CFE were responsible for funding acquisition. All authors approved the final version of the manuscript. The dataset from this study is available on the Mendeley Data platform and may be accessed through the following Doi: 10.17632/f8fhyk9ff6.1.

